# Culture and identification of a “Deltamicron” SARS-CoV-2 in a three cases cluster in southern France

**DOI:** 10.1101/2022.03.03.22271812

**Authors:** Philippe Colson, Pierre-Edouard Fournier, Jeremy Delerce, Matthieu Million, Marielle Bedotto, Linda Houhamdi, Nouara Yahi, Jeremy Bayette, Anthony Levasseur, Jacques Fantini, Didier Raoult, Bernard La Scola

## Abstract

Multiple SARS-CoV-2 variants have successively, or concommitantly spread worldwide since summer 2020. A few co-infections with different variants were reported and genetic recombinations, common among coronaviruses, were reported or suspected based on co-detection of signature mutations of different variants in a given genome. Here we report three infections in southern France with a Delta 21J/AY.4-Omicron 21K/BA.1 “Deltamicron” recombinant. The hybrid genome harbors signature mutations of the two lineages, supported by a mean sequencing depth of 1,163-1,421 reads and mean nucleotide diversity of 0.1-0.6%. It is composed of the near full-length spike gene (from codons 156-179) of an Omicron 21K/BA.1 variant in a Delta 21J/AY.4 lineage backbone. Importantly, we cultured an isolate of this recombinant and sequenced its genome. It was observed by scanning electron microscopy. As it is misidentified with current variant screening qPCR, we designed and implemented for routine diagnosis a specific duplex qPCR. Finally, structural analysis of the recombinant spike suggested its hybrid content could optimize viral binding to the host cell membrane. These findings prompt further studies of the virological, epidemiological, and clinical features of this recombinant.

## Introduction

The current SARS-CoV-2 pandemic has highlighted since the summer of 2020 the successive or concomittant emergence of numerous viral variants, each causing a specific epidemic.^1-3^ Some of these variants spread to become pandemic while others remained epidemic in a restricted geographical area. The variants characterized so far have been shaped by nucleotide substitutions, insertions or deletions. However, another major evolutionary mechanism of RNA viruses is genetic recombination, which is very common among coronaviruses.^4-8^ It requires co-infection of the same host cell by two viruses, which may be two distinct mutants or variants.^9^ Therefore, the frequency of creation of recombinants between two variants depends on the duration of their co-circulation, on the time until viral clearance, and on the number of people exposed to both viruses. Co-infections with two variants were reported including recently with SARS-CoV-2 Delta and Omicron variants.^10-13^ Furthermore, genetic recombinations were reported or suspected, based on the concurrent detection in consensus genomes of signature mutations of different mutants or variants.^10,12,14-24^ A study detected up to 1,175 (0.2%) putative recombinant genomes among 537,360 genomes and estimated that up to 5% SARS-CoV-2 having circulated in the USA and UK could be recombinants.^16^

Two pandemic variants, Delta and Omicron 21K (Nextclade classification^25,26^)/BA.1 (Pangolin classification^27^), recently succeeded each other as the predominant viruses but co-circulated for a period of several weeks, creating conditions for co-infections and subsequently recombinations. This period spanned between December 27^th^, 2021 and February 14^th^, 2022 in our geographical area, as assessed by our SARS-CoV-2 genotypic surveillance based on variant-specific qPCR and next-generation genomic sequencing.^3,28,29^ In January 2022, genomes harboring mutations from both Delta and Omicron 21K/BA.1 variants were reported in Cyprus but it was questioned whether sequences might have resulted from contamination.^23^ Still, 15 genomes as of 27/02/2022 being hybrids of these two variants and highly similar between each other were reported since February 2022 (https://github.com/cov-lineages/pango-designation/issues/444). We herein report three infections by a recombinant SARS-CoV-2 Delta 21K/AY.4-Omicron 21K/BA.1 whose genome is highly similar to the 15 previously reported genomes and the isolation of the recombinant virus from one of the patients.

## Materials and methods

Nasopharyngeal samples were collected from two patients in our university hospital institute (Méditerranée Infection; https://www.mediterranee-infection.com/) and tested for SARS-CoV-2 infection by real-time reverse transcription-PCR (qPCR) as previously described.^3,28,29^ The third patient was sampled in a private medical biology laboratory in southern France (Inovie Labosud, Montpellier, France). qPCR assays that screen for SARS-CoV-2 variants were performed as recommended by French public health authorities (https://www.data.gouv.fr/fr/datasets/donnees-de-laboratoires-pour-le-depistage-indicateurs-sur-les-mutations/). In our center this included the detection of spike mutation among which K417N (Thermo Fisher Scientific, Waltham, USA), combined with testing with the TaqPath COVID-19 kit (Thermo Fisher Scientific) that target viral genes ORF1, N (nucleocapsid) and S (spike), as previously reported.^3,28,29^ The private medical laboratory used the ID SARS-CoV-2/VOC Revolution Pentaplex assay (ID Solutions, Grabels, France) that detects spike mutations K417N, L452R, and E484K (Pentaplex assay, ID Solution, France).

SARS-CoV-2 genomes were sequenced in the framework of genomic surveillance implemented since February 2020^3^ in our institute. Next-generation sequencing was performed with the Illumina COVID-seq protocol on the NovaSeq 6000 instrument (Illumina Inc., San Diego, CA, USA) or with the Oxford Nanopore technology (ONT) on a GridION instrument (Oxford Nanopore Technologies Ltd., Oxford, UK) combined with prior multiplex PCR amplification according to the ARTIC procedure (https://artic.network/), as previously described,^3,28^ with the ARTIC nCoV-2019 Amplicon Panel v4.1 of primers (IDT, Coralville, IA, USA). Then, sequence read processing and genome analysis were performed as previously described.^3,28^ Briefly, for Illumina NovaSeq reads, base calling was performed with the Dragen Bcl Convert pipeline [v3.9.3; https://emea.support.illumina.com/sequencing/sequencing_software/bcl-convert.html (Illumina Inc.)], mapping was performed with the bwa-mem2 tool (v. 2.2.1; https://github.com/bwa-mem2/bwa-mem2) on the Wuhan-Hu-1 isolate genome (GenBank accession no. NC_045512.2) then cleaned with Samtools (v. 1.13; https://www.htslib.org/), variant calling was carried out with freebayes (v. 1.3.5; https://github.com/freebayes/freebayes) and consensus genomes were built using Bcftools (v. 1.13; https://samtools.github.io/bcftools/bcftools.html). ONT reads were processed with the ARTIC-nCoV-bioinformaticsSOP pipeline v1.1.0 (https://github.com/artic-network/fieldbioinformatics). Nucleotide and amino acid changes relatively to the Wuhan-Hu-1 isolate genome were obtained using the Nextclade tool (https://clades.nextstrain.org/).^25,26^ Nextstrain clades and Pangolin lineages were determined using the Nextclade web application (https://clades.nextstrain.org/)^25,26^ and the Pangolin tool (https://cov-lineages.org/pangolin.html),^27^ respectively. Genome sequences described here were deposited in the GenBank sequence database (https://www.ncbi.nlm.nih.gov/genbank/)^30^ (OM990851, OM990852, OM991095, OM991295), and on the IHU Méditerranée Infection website (https://www.mediterranee-infection.com/sars-cov-2-recombinant/). The Simplot software (https://sray.med.som.jhmi.edu/SCRoftware/SimPlot/)^31^ was used for recombination analysis. Phylogeny was reconstructed by the IQTree (v2.1.3; http://www.iqtree.org/)^32^ or MEGA X^33^ (v10.2.5; https://www.megasoftware.net/) tools and visualized with MEGA X after sequence alignment with MAFFT (https://mafft.cbrc.jp/alignment/server/).^34^ As phylogenetic analysis can hardly applies to sequences that have different evolutionary histories, we built two separate trees, a first one for the regions classified as of the Delta 21J/AY.4 variant (positions 1-22,128 and 25,519-29,903 in reference to the genome of the Wuhan-Hu-1 isolate), and a second one for the regions classified as of the Omicron 21K/BA.1 variant (positions 22,129-25,519). The 10 genomes the closest genetically to these fragments of the genome obtained here were selected through a BLAST^35^ search among genomes of the Delta 21J/AY.4 and Omicron 21K/BA.1 variants in the sequence database of our institute that contains approximately 50,000 SARS-CoV-2 genomes; then, they were incorporated in the phylogeny together with the genome of the Wuhan-Hu-1 isolate. As a matter of fact, it should not be acceptable to make a phylogenetic tree based on concatenation of sequences from different origin.

SARS-CoV-2 culture isolation was performed by inoculating 200 µL of respiratory sample on Vero E6 cells as previously described.^36^ Cytopathic effect was observed by inverted microscopy. Viral particles were visualized in the culture supernatant by scanning electron microscopy with a SU5000 microscope (Hitachi High-Technologies Corporation, Tokyo, Japan), as previously described.^37^

Structural predictions of the spike protein were performed as previously described.^28,38,39^ Briefly, amino acid changes were introduced in the framework of a complete 14-1,200 structure of the original SARS-CoV-2 20B spike and missing amino acids were incorporated with the Robetta protein structure prediction tool (https://robetta.bakerlab.org/) before energy minimization through the Polak-Ribière algorithm.

A in house duplex qPCR assay specific of the SARS-CoV-2 recombinant was designed that targets the genomes of the Delta 21J [targeted mutation: A11201G in the Nsp6 gene (corresponding to amino acid substitution T77A)] and the Omicron 21K/BA.1 [targeted mutations: A23040G, G23048A, A23055G in the spike gene (Q493R, G496S, Q498R)] variants. The sequences of the primers and probes (in 5’-3’ orientation) are as follows: (i) for the Delta 21J-targeting system: forward primer, CTGCTTTTGCAATGATGTTTGT; reverse primer, TACGCATCACCCAACTAGCA; probe, 6FAM-CTTGCCGCTGTAGCTTATTTTAAT (primers and probe concentrations in the mix were 200 nM and 150 nM, respectively); (ii) for the Omicron 21K/BA.1-targeting system: forward primer, CCTTGTAATGGTGTTGAAGGTTTT; reverse primer, CTGGTGCATGTAGAAGTTCAAAAG; probe, 6VIC-TTTACGATCATATAGTTTCCGACCC (primers and probe concentrations in the mix were 250 nM and 200 nM, respectively).

This study was approved by the ethics committee of University Hospital Institute Méditerranée Infection (No. 2022-001).

## Results

The three case-patients were SARS-CoV-2-diagnosed on nasopharyngeal samples collected in February 2022. Cycle threshold values (Ct) of diagnostic qPCR were between 20-21. The patients were below 40 years of age. They resided in southern France and did not travel abroad recently. They presented mild respiratory symptoms. Two were vaccinated against SARS-CoV-2 (with two or three doses administered). Variant screening qPCR for the 2 samples collected in our institute showed positivity for the K417N mutation while the TaqPath COVID-19 kit provided positive signals for all three genes targeted (ORF1, S, and N). The third sample showed positivity for the K417N mutation and negativity for the L452R and E484K mutations. Thus, overall, qPCR carried out on the three samples were indicative of a Omicron variant.

The three viral genomes [GenBank Accession no. OM990851, OM990852, OM991095 (https://www.ncbi.nlm.nih.gov/genbank/)^30^; available on the IHU Méditerranée Infection website (https://www.mediterranee-infection.com/sars-cov-2-recombinant/)] were hybrids of Delta 21J/AY.4 and Omicron 21K/BA.1 variant genomes (**Figures 1a, b, c; Table 1**). At positions harboring mutations compared to the genome of the Wuhan-Hu-1 isolate, mean sequencing depth was between 1,163-1,421 reads and mean prevalence of the majoritary nucleotide was between 99.4-99.9% (with minimum values between 80.3-98.7%), ruling out the concurrent presence of two variants in the samples either due to co-infection or to contamination. In this SARS-CoV-2 recombinant, most of the spike gene was replaced in a Delta 21J/AY.4 matrix by an Omicron 21K/BA.1 sequence (**Figures 1a, b, c**). Indeed, the recombination sites were located between nucleotide positions 22,034 and 22,194 for the first one, and between nucleotide positions 25,469 and 25,584 for the second one. These regions corresponds to amino acids 158 to 211 of the spike protein and to amino acids 26 to 64 of the ORF3a protein whose gene is contiguous to the spike gene. The Simplot recombination analysis tool provided congruent results.

**Table 1.**
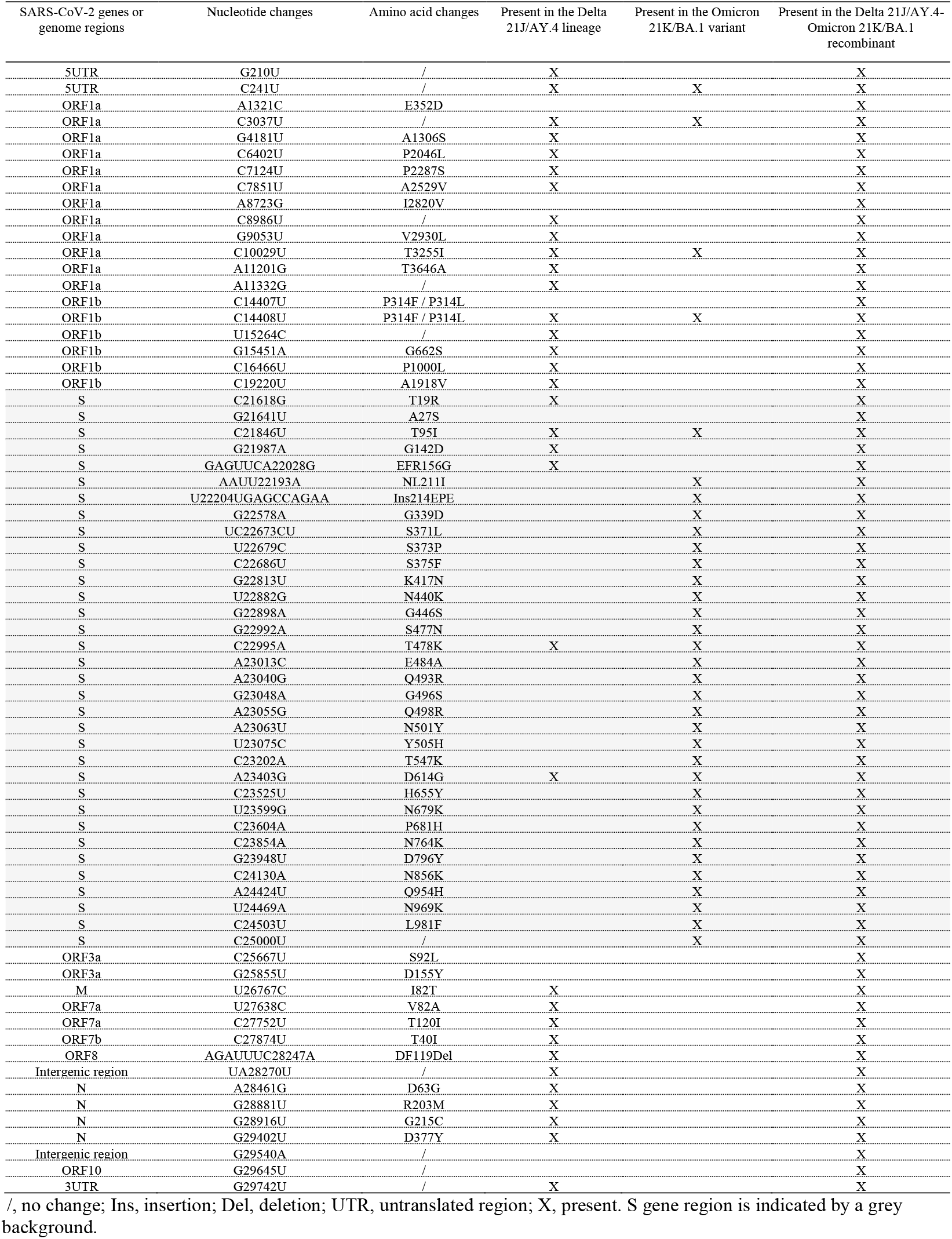
Nucleotide and amino acid changes in the Deltamicron recombinant according to their presence/absence in the Delta 21J/AY.4 lineage and the Omicron 21K/BA.1 variant.

**Figure 1.**
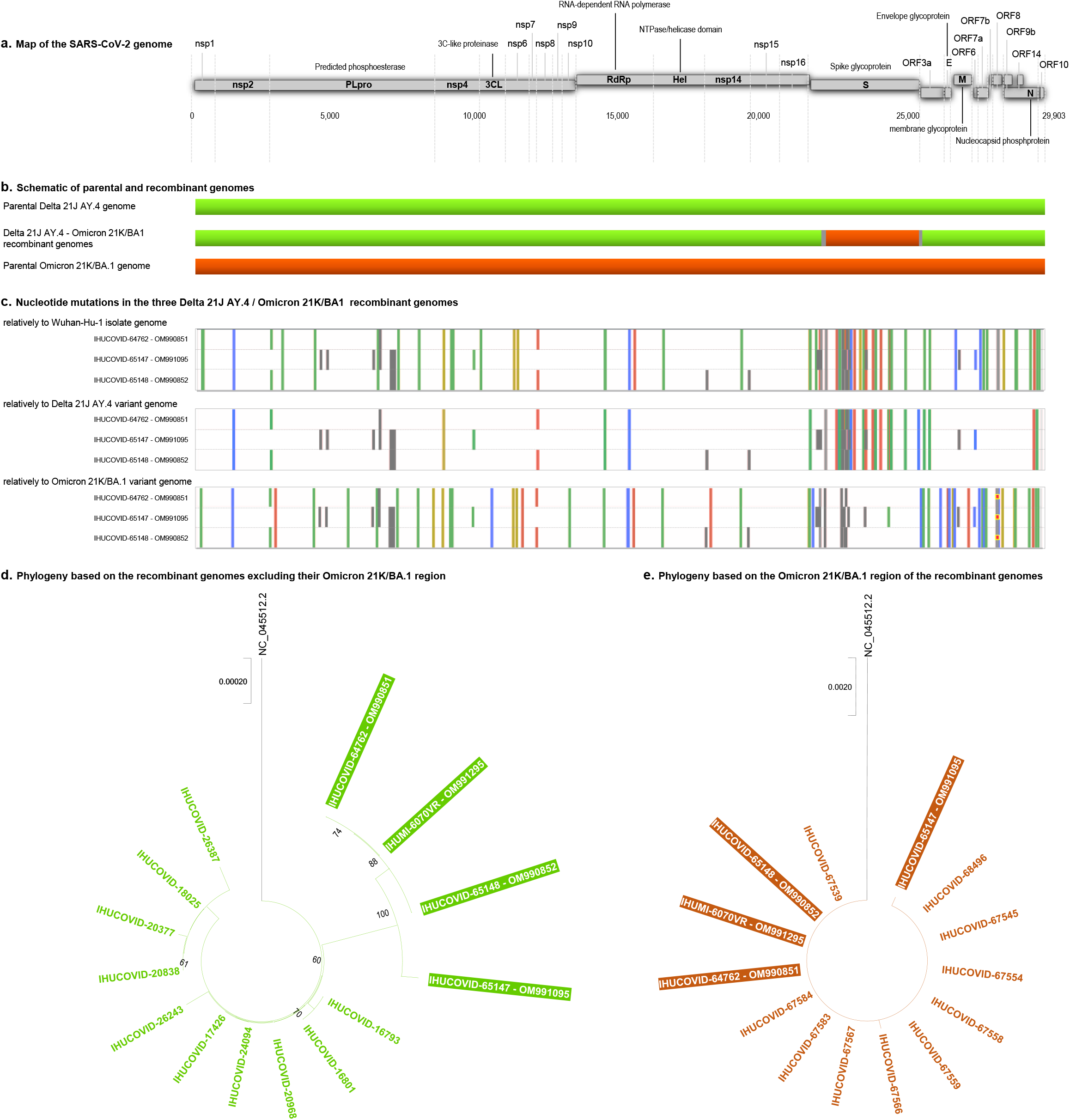
Schematic of the SARS-CoV-2 Delta 21J/AY.4-Omicron 21K/BA.1 recombinant genome, and phylogenetic trees based on different regions of the Deltamicron genome reflecting the origin of most of the viral genome from a Delta 21J/AY.4 variant and the origin of a region spanning a large part of the spike gene from a Omicron 21K/BA.1 variant. a: Map of the SARS-CoV-2 genome. b: Schematic representation of parental and recombinant genomes. c: Mutations in the three Delta 21J/AY.4-Omicron 21K/BA1 recombinant genomes. Adapted from screenshots of the nextclade web application output (https://clades.nextstrain.org).^25,26^ Color codes for nucleotide mutations and are marked regions as follows: Green: U; yellow: G; blue: C; red: A; grey: deletions or uncovered regions. Genomes are labelled with the identifiers of the IHU Méditerranée Infection and the GenBank (https://www.ncbi.nlm.nih.gov/genbank/)^30^ sequence databases. d, e: Phylogenetic trees based on different regions of the Deltamicron genomes showing the origin of most of the viral genome from a Delta 21J/AY.4 variant (d) and the origin of a region spanning a large part of the spike gene from a Omicron 21K/BA.1 variant (e). The 10 genomes the most similar genetically to these regions of the recombinant genomes obtained here were selected from the sequence database of our institute through a BLAST^35^ search, then were incorporated in the phylogeny together with the genome of the Wuhan-Hu-1 isolate. Sequences are labelled with the identifiers of the IHU Méditerranée Infection and the GenBank (https://www.ncbi.nlm.nih.gov/genbank/)^30^ sequence databases.

Genomes from two of the three patients were identical and clustered in the phylogenetic analysis (**Figures 1d, e**) despite no epidemiological link was documented between these two patients. The third genome exhibited 5 nucleotide differences. Phylogenetic analyses showed that most of the recombinant genomes was most closely related to Delta 21J/AY.4 variant genomes identified in our institute, while the region spanning a large part of their spike gene was most closely related to Omicron 21K/BA.1 variant genomes identified in our institute.

The respiratory sample from which the first recombinant genome was obtained was inoculated on Vero E6 cells the day following recombinant identification, and cytopathic effect was observed after 4 days (**Figures 2a, b**; collection of strains of IHU Méditerranée Infection, no. IHUMI-6070VR). The same day, supernatant was collected and next-generation genome sequencing was performed using Nanopore technology on a GridION instrument after PCR amplification with Artic primers, which allowed obtaining the genome sequence of the viral isolate 8 h later [GenBank Accession no. OM991295 (https://www.ncbi.nlm.nih.gov/genbank/)^30^; available on the IHU Méditerranée Infection website (https://www.mediterranee-infection.com/sars-cov-2-recombinant/)]. At mutated positions compared to the Wuhan-Hu-1 isolate genome, mean sequencing depth was 2,771 reads and mean prevalence of the majoritary nucleotide was 99.1% (minimum, 95.1%), and the consensus genome was identical to that obtained from the respiratory sample, showing unambiguously that the virus isolated was the Delta 21J/AY.4-Omicron 21K/BA.1 recombinant. Finally, viral particles were observed in the culture supernatant by scanning electron microscopy with a SU5000 microscope within minutes after supernatant collection (**Figure 2c**).

**Figure 2.**
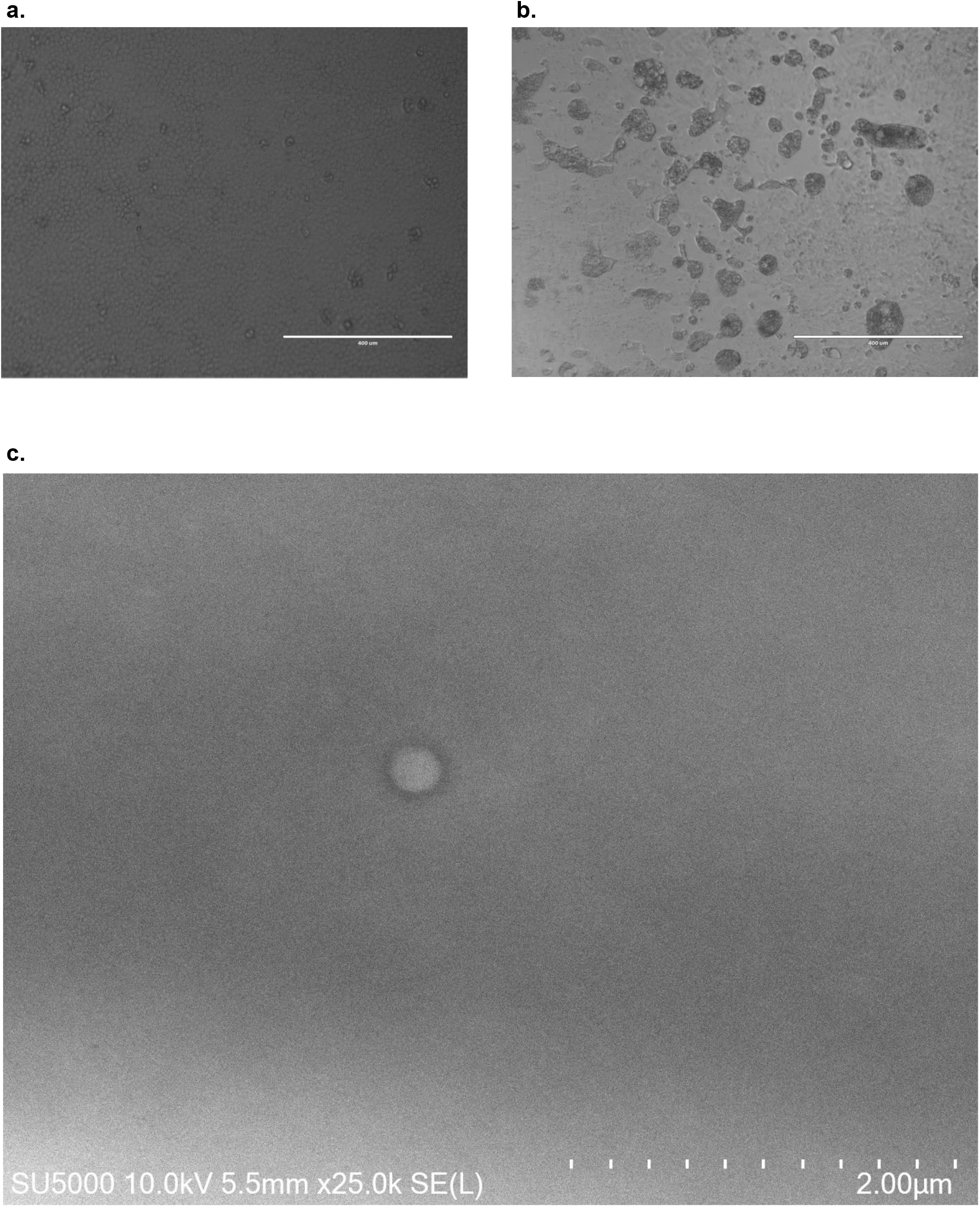
Microscopy images of the virus cytopathic effect (a, b) and of a viral particle (c) in culture of the SARS-CoV-2 Delta 21J/AY.4-Omicron 21K/BA.1 recombinant on Vero E6 cells. a. Absence of cytopathic effect (negative control: absence of virus); b. Cytopathic effect observed 4 days post-inoculation on Vero E6 cells of the respiratory sample of the first patient for whom the SARS-CoV-2 Delta 21J/AY.4-Omicron 21K/BA.1 recombinant was identified. c. Scanning electron microscopy image was obtained with a SU5000 microscope (Hitachi High-Technologies Corporation, Tokyo, Japan).

The overall structure of the recombinant spike protein was predicted (**Figures 3a-c)**. When superimposed with the spike of the Omicron 21K/BA.1 variant, the main structural changes were located in the N-terminal domain (NTD). In this region, the surface of the recombinant spike protein is enlarged, flattened, and more electropositive, a property that is characteristic of the Delta 21J/AY.4 NTD (**Figure 3c**). In the initial interaction of the virus with the plasma membrane of the host cells, the NTD is attracted by lipid rafts, which provides an electronegative landing platforms for the spike.^38,39^ Thus, an increase in the electrostatic surface of the NTD is expected to accelerate the binding of the virus to lipid rafts, which may confer a selective kinetic advantage against virus competitors.^38^ The receptor binding domain (RBD) of the recombinant is clearly inherited from the Omicron 21K/BA.1 variant (**Figure 3c**). The consequence is also an increase in the electrostatic surface potential of the RBD, which may facilitate the interaction with the electronegative interface of the ACE-2 cellular receptor. Overall, this structural analysis suggests that the recombinant virus could have been selected on the basis of kinetic properties conferred by a convergent increase of the electrostatic potential of both the NTD and the RBD, together with an enlargement of the NTD surface, all features that suggest an optimization of virus binding to the host cell membrane.

**Figure 3.**
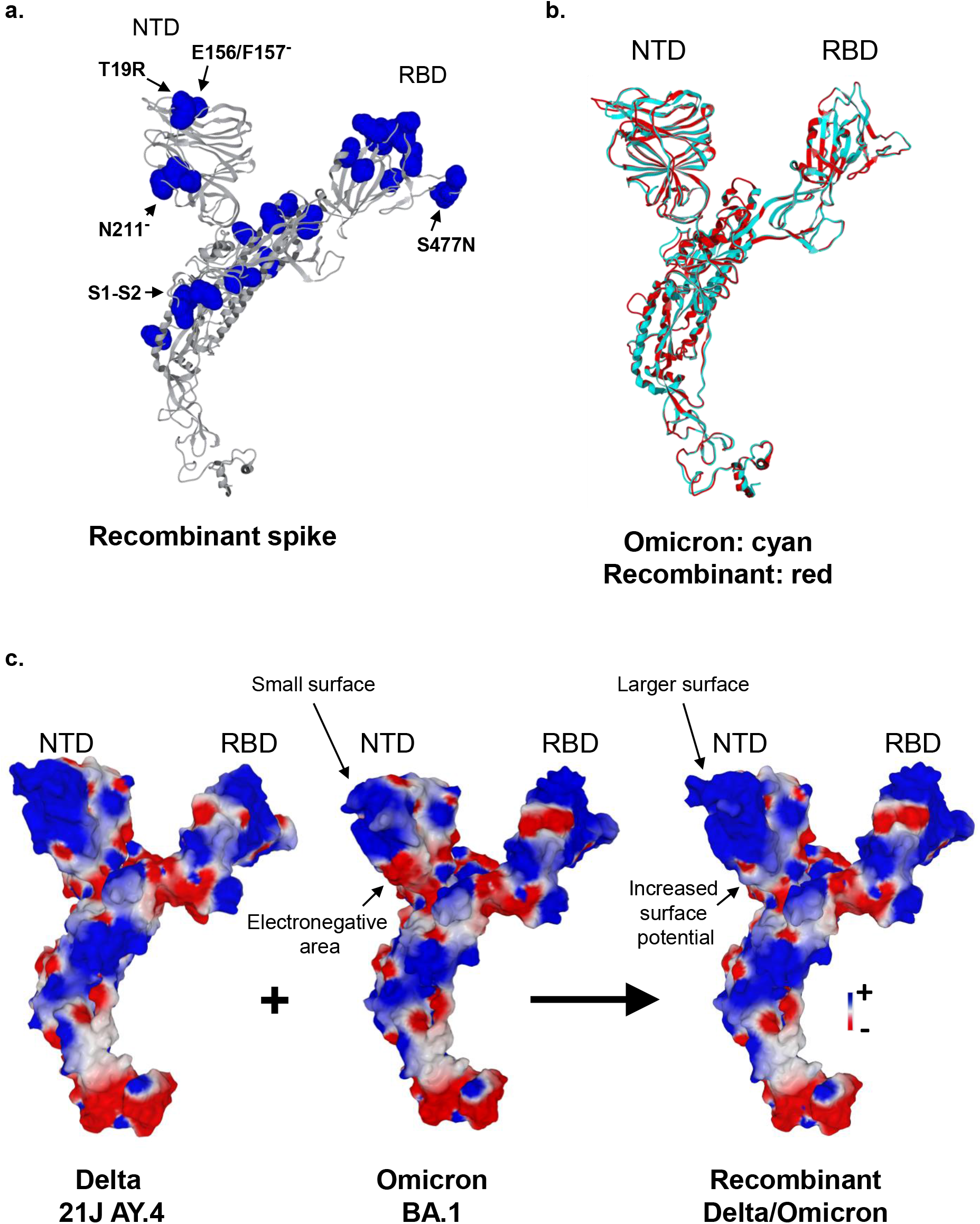
Schematic of the predicted structure of the spike protein of the SARS-CoV-2 Delta 21J/AY.4-Omicron 21K/BA.1 recombinant. a. Overall structure of the recombinant spike protein. The secondary structure is in grey, mutated amino acids are in blue. NTD, N-terminal domain; RBD, receptor binding domain; S1-S2, cleavage site. b. Superimposition of the secondary structure of the Omicron 21K/BA.1 variant (in cyan) and recombinant (in red) spike proteins. NTD, N-terminal domain; RBD, receptor binding domain. c. Comparison of the electrostatic surface potential of the spike proteins in Delta 21J/AY.4 lineage, Omicron 21K/BA.1 variant, and in the recombinant. The color scale (negative in red, positive in blue, neutral in white) is indicated. NTD, N-terminal domain; RBD, receptor binding domain.

Currently used qPCR screening assays were not able to discriminate this Delta 21J/AY.4-Omicron 21K/BA.1 recombinant and the Omicron 21K/BA.1 variant because the Delta variant signature mutation detected is absent from the recombinant genome. Therefore, we attempted to promptly implement a specific qPCR assay that could be used to detect the recombinant for routine diagnosis use. This was achieved in one day by selecting qPCR systems from our toolbox of dozens of in house qPCR systems that were designed since the emergence of the first variant during summer 2020 to specifically target SARS-CoV-2 variants or mutants.^3,40^ Two systems that target either the Delta 21J variant or the Omicron BA.1 variant were combined in a duplex qPCR that can screen for the recombinant. In preliminary assessment, both tested recombinant-positive samples were positive for the Delta 21J and Omicron BA.1 targets. In addition, 7 Delta non-21J-positive samples were negative for both targets, 4 Delta 21J-positive samples were positive for the Delta 21J target but negative for the Omicron BA.1 target, and 3 Omicron BA.1-positive samples were negative for the Delta 21J target but positive for the Omicron BA.1 target.

## Discussion

It is increasingly demonstrated that the genomes of most biological entities, whatever their level of complexity, are mosaics of sequences from various origins.^41-45^ The present observations show us in real life the recombination potential of SARS-CoV-2, already largely established for other coronaviruses.^4-6,8,46^ and reported or suspected for SARS-CoV-2.^10,12,14-24^ SARS-CoV-2 recombinations were difficult to spot when only genetically very similar viruses were circulating, as was the case in Europe during the first epidemic episode with mutants derived from the Wuhan-Hu-1 virus. The increasing genetic diversity of SARS-CoV-2, the tremendous number of infections at global and national levels, and the unprecedented global effort of genomic sequencing (https://covariants.org/per-country),^3,25,47^ raised the probability of detecting recombinants. Such observations will probably make it possible in the short or medium term to assess the recombination rate of SARS-CoV-2, whether there are recombination hotspots, and to what extent recombinations between different variants can generate new viable, and epidemic variants. This questions on the impact of recombinations on viral replication and transmissibility, and on clinical severity, as well as on the virus ability to escape neutralizing antibodies elicited by vaccines or a previous infection. In this view, culture isolation of SARS-CoV-2 recombinants as was carried out here for the first time to our knowledge is of primary importance. This variant mentioned recently has no known epidemic potential. This will allow studying their phenotypic properties, among which their replicative capacity in various cell lines, their sensitivity to antibodies, or their genetic evolution *in vitro*. Concurrently, a high level of genomic surveillance must be maintained in order to detect and characterize all recombination events and circulating recombinants, which is a critical scientific and public health issue.

## Data Availability

The dataset generated during the current study is available from the GenBank database (https://www.ncbi.nlm.nih.gov/genbank/) (GenBank Accession no. OM990851, OM990852, OM991095, OM991295) and from the IHU Mediterranee Infection website (https://www.mediterranee-infection.com/sars-cov-2-recombinant/).

https://www.mediterranee-infection.com/sars-cov-2-recombinant/

## Acknowledgments

We are very grateful to Clio Grimaldier, Rita Zgheib, Claudia Andrieu, Ludivine Bréchard, Raphael Tola, Anthony Fontanini, and Jacques Bou Khalil for their technical help.

## Author contributions

Study conception and design: Philippe Colson, Pierre-Edouard Fournier, Jacques Fantini, Didier Raoult, Bernard La Scola. Materials, data and analysis tools: Philippe Colson, Pierre-Edouard Fournier, Jeremy Delerce, Matthieu Million, Marielle Bedotto, Linda Houhamdi, Nouara Yahi, Jeremy Bayette, Jacques Fantini. Data analyses: Philippe Colson, Pierre-Edouard Fournier, Jeremy Delerce, Marielle Bedotto, Anthony Levasseur, Jacques Fantini, Didier Raoult, Bernard La Scola. Writing of the first draft of the manuscript: Philippe Colson, Jacques Fantini, and Pierre-Edouard Fournier. All authors read, commented on, and approved the final manuscript.

## Data availability

The dataset generated during the current study is available from the GenBank database (https://www.ncbi.nlm.nih.gov/genbank/)^30^ (GenBank Accession no. OM990851, OM990852, OM991095, OM991295) and from the IHU Méditerranée Infection website (https://www.mediterranee-infection.com/sars-cov-2-recombinant/).

## Ethics

This study has been approved by the ethics committee of the University Hospital Institute Méditerranée Infection (No. 2022-001). Access to the patients’ biological and registry data issued from the hospital information system was approved by the data protection committee of Assistance Publique-Hôpitaux de Marseille (APHM) and was recorded in the European General Data Protection Regulation registry under number RGPD/APHM 2019-73.

